# Large language model-assisted causal machine learning for identifying fatigue-related poor glycated hemoglobin in type 2 diabetes

**DOI:** 10.1101/2025.02.10.25321977

**Authors:** Herdiantri Sufriyana, Debby Syahru Romadlon, Rudy Kurniawan, Safiruddin Al Baqi, Emmanuel Ekpor, Eric Peprah Osei, Hsiao-Yean Chiu, Emily Chia-Yu Su

## Abstract

**Background:** Fatigue is common but mostly untreated in type 2 diabetes, since it requires a diagnostic workup which is hardly justified by fatigue alone. Individually identifying fatigue-related cause help decide effective follow-up and prioritize resource utilization. This study aimed to determine whether glycated hemoglobin (HbA1c) level contributes to fatigue for each individual case using a fatigue prediction model based on large language model (LLM)-assisted structural causal modeling (SCM).

**Methods:** Retrospective cohort design was applied to collect data from diabetes management centers in Indonesia. We conducted SCM to select predictors among HbA1c and other variables for fatigue predictive modeling. Causal diagram was constructed by inferring the causal direction for each pair of correlated variables via LLM, i.e., GPT-4. The models were trained using eight machine learning (ML) algorithms. The best one was selected among the models that fulfilled sufficient sample size, was well-calibrated, and had positive net benefit using threshold closest to 95% specificity in our data. We chose the best model based on the area under curve (AUC) of receiver operating characteristics (ROC) and the concordance between the feature impact on model output based on the beeswarm plot of the Shapley additive explanation (SHAP) values and the effect size based on SCM. The SHAP waterfall plot was utilized to quantify HbA1c contribution to fatigue for each individual case.

**Results:** Individuals with type 2 diabetes receiving OHA last 3 months (*n*=281) were more likely to report fatigue when they had poor HbA1c (adjusted odds ratio [aOR]=6.1, 95% CI 2.5, 14.4), comorbidity (aOR=25.2, 95% CI 12.1, 52.4), or a need for insulin treatment (aOR=3.6, 95% CI 2.0, 6.4). The best model used random forest algorithm (AUC-ROC=0.966, 95% CI 0.962, 0.969). Fatigue-related poor HbA1c could be individually identified among 95.1% (95% CI 94.5, 95.7) of those who reported fatigue.

**Conclusions:** We have developed a web application and nomogram for identifying fatigue-related poor HbA1c for each individual case. Future studies are warranted for external validation and randomized trials to examined the validity and impact of the cause identifier of fatigue in this study.

## Introduction

Pooled prevalence of fatigue was 50% of individuals with type 2 diabetes [1, 2]. Fatigue influences their overall well-being and ability to manage their condition [3]. Fatigue in type 2 diabetes was correlated to the reduction in quality of life and functional status [4]. Meanwhile, 62% of such individuals were never or rarely treated [2]. Untreated fatigue can undermine diabetes self-management (DSM) [5] while DSM education may reduce HbA1c level [6]. Individuals with slightly higher HbA1c level (6.5% to <7.0% vs. <6.5%) within a year was 1.2 times more likely to have diabetic eye, lower-extremity amputation, or cerebrovascular/heart/renal diseases. An unmet need for fatigue intervention is possibly because deciding the intervention requires a diagnostic workup which burdens healthcare costs and may lead to resource overutilization. Such decision is hardly justified by self-reported fatigue alone. Therefore, identifying fatigue-related poor HbA1c at individual level would help decide the effective follow-up and prioritize resource utilization.

Fatigue may be resulted from poor glycated hemoglobin (HbA1c) [7] or other factors [1, 7-13]. They included physiological (e.g., diabetes treatment [7] and duration [7, 8], comorbidity [9-13]) and psychological factors (e.g., depression [1, 8]). The aforementioned factors might be confounded by sociodemographic factors (e.g., age, sex, marital and educational status) [7, 8] and behavior and lifestyle (e.g., sleep quality [7, 14], cigarette smoking [14-16]). Identifying a fatigue-related cause help avoid delay in effective intervention and prioritizes resource utilization. Such identification is straightforward in a medical condition with a single factor, such as infectious diseases, and it helps decide a targeted intervention leading to symptom alleviation, immediately. However, identifying a symptom-related cause has never been conducted at individual level for a multifactorial condition in medicine, such as fatigue among individuals with type 2 diabetes.

The previous studies [6, 7] implied a bidirectional relationship between HbA1c and fatigue. Nonetheless, a causal identification is well-accepted for HbA1c rise but not fatigue; hence, it requires an efficient diagnostic workup. Ideally, we need to quantify the impact of each cause on fatigue, then select or prioritize that with the highest impact as the primary cause in order to effectively alleviate fatigue at individual level, which refers to specific causality [17]. It is approached by logic-based causality [18] using dichotomy (no/yes) which is ambiguous in medical conditions that have multiple causes. Meanwhile, a causal inference in medicine is conducted at population level, e.g., examining whether a drug is effective to cure a disease [19], so called general causality. It is approached by covariance-based causal analysis [20, 21]. However, there was no previous study that applies covariance-based causal analysis to approach specific causality, including to identify whether poor HbA1c is related to fatigue for each individual case.

Over the past three decades, causal inference has become feasible not only through a randomized-controlled trials (RCT) but also observational studies [22, 23]. Both approaches rely on covariance-based causal analysis [21, 24]. However, observational studies have been enabled to specific causality by recent advancements in artificial intelligence (AI), particularly in structural causal modeling (SCM) and machine learning (ML) explainability [25, 26]. Explainable AI allows covariance-based, specific causality using an ML prediction model with two criteria: (1) it is accurate; and (2) the way it results in the prediction (i.e., explainability) resembles causal structure as inferred by SCM. The challenge is that the hypothetical effect direction between variables in causal diagram, as a prerequisite for SCM, cannot be inferred using data but domain knowledge [25]. AI progress in the last two years scaled up such knowledge-driven analysis using large language model (LLM) such as ChatGPT [27]. Unlike a human individual, LLM also allows inquiry an answer to a specific question from an internet-size knowledge base.

This study aimed to determine whether HbA1c level contributes to fatigue for each individual case using a fatigue prediction model based on LLM-assisted SCM.

## Methods

### Study design and setting

We applied retrospective cohort design to collect data between January and June 2024. The settings were diabetes management centers in Indonesia. The eligibility criterion of participants in this study was individuals with type 2 diabetes. This study was approved by the Joint Institutional Review Board of the Ethical Committee of Medical Research at the Faculty of Dentistry, Universitas Jember (no.: 2683/UN25.8/KEPK/DL/2024).

To achieve the objective, we need to conduct both causal and predictive modeling. The former was intended to select candidate predictors for the latter. For predictive modeling, we developed a prognostic prediction model with cross validation. The predictor information was obtained from the past 3 months to the time of prediction, while the outcome information was asked at the time of prediction. The best model was selected to achieve high sensitivity and precision/positive predictive value (PPV) with the feature importance similar to their effect sizes in the causal modeling. Our model was intended to identify the impact of HbA1c, as the predictor, on the model output, i.e., the predicted outcome of fatigue, among individuals with the true outcome of fatigue.

### Data collection and preprocessing

We collected data for 10 variables by interviewing the participants at the time of survey and reviewing their medical records from the past 3 months. At the time of survey, we asked the participants to obtain 6 variables: (1) age (years); (2) sex (female/male); (3) marital status (unmarried/married/widowed or divorced); (4) educational status (no education/primary school/junior secondary school/senior secondary school/college graduate); (5) cigarette smoking (no/yes); (6) fatigue (no/yes). The remaining 4 variables were obtained from medical records: (1) HbA1c (%); (2) diabetes treatment (OHA/OHA + insulin); (3) diabetes duration (years); and (4) comorbidity (no/yes).

We preprocessed data for numerical variables to achieve normal distribution and remove outliers. All numerical variables were assessed for normality (see “Statistical analysis”). We transformed variables using either simple or Box-Cox transformation and reassessed their normality. If the normal distribution could not be achieved for a variable, then it was categorized based on domain knowledge. The categorized version was used for causal modeling, but the numerical one was retained for predictive modeling. Subsequently, we removed outliers of the remaining numerical variables (see “Statistical analysis”) by assigning missing values. We also added a missingness variable (non-missing/missing) for a variable with missing values.

To impute missing values, we applied multiple imputation by chained equation (MICE) using predictive mean matching methods with 10-time imputation. We utilized the correlation matrix (see “Structural causal modeling”) to identify which variables were correlated to a missingness variable. Such variables, if any, were used to impute the corresponding variable with missing values; otherwise, the variable was imputed by assuming it as missing completely at random.

### Structural causal modeling

Causal modeling was conducted using structural causal modeling (SCM) method. In this method, domain knowledge is employed to hypothetically construct a causal diagram, while data are used to estimate the causal effect. To simplify SCM procedures for constructing a causal diagram, first, we used a statistical test to identify a pair of variables that were correlated, then, we determined the causal direction using LLM. Subsequently, we applied multivariate regression analysis to estimate the effect of a variable that hypothetically caused fatigue.

We inferred a correlation between two variables for all possible combinations using a statistical test to construct a correlation matrix. The details of the statistical tests are specified (Figure S1). To correct the multiple testing effects, the Benjamini-Hochberg method was applied. We determined the causal direction for a pair of variables if they were significantly correlated (see “Statistical analysis”).

To determine the causal direction, we used LLM, i.e., GPT-4 via ChatGPT from August 16 to 26, 2024. A previous study has engineered a specific prompt for asking a causal direction [27]. We applied this prompt format with a definition for each variable being prompted, as shown below.

> *“You are a helpful assistant for causal reasoning. Which cause-and-effect relationship is more likely?*
>
> *A. changing **Variable 1** causes a change in **Variable 2**.*
>
> *B. changing **Variable 2** causes a change in **Variable 1**.*
>
> *Definitions:*
>
> ***Variable 1****: Definition 1.*
>
> ***Variable 2****: Definition 2.*
>
> *Let’s work this out in a step-by-step way to be sure that we have the right answer.*
>
> *Then provide your final answer within the tags A/B.”*

The causal diagram was used to determine covariates (*Z*) in a multivariate regression analysis between an independent variable (*X*) and fatigue as dependent variable (*Y*). We only included variables *L* as the covariates *L* if they were confounders or mediators. A variable *L* was considered a confounder if it was correlated to both X and Y with *L* ➔ *X* and *L* ➔ *Y* causal directions, respectively. Meanwhile, a variable *L* was considered a mediator if it was correlated to both X and Y with *X* ➔ *L* and *L* ➔ *Y* causal directions, respectively. We selected a variable as a candidate predictor was significantly correlated to fatigue after adjustment for the covariates (see “Statistical analysis”). This means that the variable has a significant effect on fatigue beyond the mediator effect if any.

### Predictive modeling

We developed a prediction model using candidate predictors as selected by SCM. Several models were developed 8 machine learning (ML) algorithms, including deep learning: (1) ridge regression (RR); (2) naïve Bayes (NB); (3) support vector machine (SVM) with linear kernel; (4) *k*-nearest neighbor (KNN); (5) decision tree (DT); (6) random forest (RF); (7) gradient boosting machine (GBM); and (8) deep neural network (DNN). We chose the former 4 algorithms because they generally require smaller sample size for model training. Nonetheless, we conducted sample size estimation for all the algorithms. In general, the hyperparameter grid for each algorithm was determined to retain all the candidate predictors and to avoid redundant approaches among the algorithms. The reasoning for the algorithm of choices and their hyperparameters are described (Tables S1 and S2).

The sample size estimation for an ML algorithm was conducted by evaluating the discrimination ability across increasing numbers of sample sizes, similar to a previous study [28]. We randomly subset our dataset from 10% to 90% by 10% increment for this evaluation. Our dataset was sufficient for an algorithm if the discrimination ability increased at a rate negatively proportional to the number of minority class of outcome divided by the number of candidate predictors (see “Statistical analysis”).

Model validation was conducted by cross validation. We applied 5-fold cross validation for hyperparameter tuning for each algorithm. Meanwhile, bootstrapping for 30 times was applied for evaluating the model with the best hyperparameters among all the ML algorithms.

The best model was selected among the eligible models. A model was eligible for selection if it fulfilled sufficient sample size estimation, was well-calibrated, and had positive net benefit using threshold close to 95% specificity in our data. To select the best model, the criteria included the discrimination ability and the concordance between the feature impact on model output and the causal effect size (see “Statistical analysis”). We determined our priority between the two criteria *post hoc* to consider tradeoff between performance in predicting the outcome and the correctness in inferring the cause. The former was intended to increase the number of eligible individuals to use the model, while the latter was intended to identify a variable that contributed the most to fatigue for each individual case, including poor HbA1c.

### Model usage

The best model was deployed via a web application. We also created a nomogram if the predictors included maximum 1 numerical variable based on a method which was previously developed for constructing an explainable nomogram for any machine learning algorithms [29]. Our model would only be used if an individual fulfills the eligibility criteria of participants in this study, similar to the sample characteristics, and reports fatigue. Furthermore, our model would only identify whether current fatigue is related to poor HbA1c last 3 months, if the predicted outcome is fatigue, i.e., matching the true outcome. We pre-computed the Shapley additive explanation (SHAP) values of the predictors to show the SHAP waterfall plot for the individual. If the SHAP value of HbA1c is positive and the highest one compared to those of other predictors, then current fatigue is related to poor HbA1c.

### Statistical analysis

We applied several statistical tests during data preprocessing and structural causal modeling. All numerical variables were assessed for normality via quantile-to-quantile plots and Shapiro-Wilk normality tests. An outlier was assigned to a missing value if it was less or greater than 1.5 interquartile range from quartile 1 and 3, respectively, of the values for each variable. In the correlation matrix, a pair of variables was significantly correlated if the false discovery rate was ≤0.05. Meanwhile, in a multivariate regression analysis, we applied logistic regression and a variable was significantly correlated to fatigue if the *p*-value was ≤0.05 after adjustment for the covariates if any.

For predictive modeling, we evaluated the models for algorithm-wise sample size sufficiency, calibration, clinical utility, discrimination ability, and model explainability and interpretability. Our dataset was sufficient for an algorithm by examining the assumption (see “Predictive modeling”) by examining how fit the rate of the discrimination ability pattern with theoretical increment based on the modified exponential decay function. For the remaining evaluation, we used point and interval estimates (i.e., 95% confidence interval, CI) by bootstrapping for 30 times. Model calibration was evaluated by fitting a linear regression of the true and predicted probabilities. A model was well-calibrated if the interval estimates of intercept and slope included 0 and 1, respectively. Brier score was also computed to identify the best calibrated model. Meanwhile, the clinical utility was evaluated by decision curve analysis, i.e., plotting the net benefit for each threshold, including that closest to 95% specificity in our data. Net benefit was positive if it was higher than those of predicting all as positives (treat all) or negatives (treat none). To evaluate discrimination ability, we computed the area under curve (AUC) of receiver operating characteristics (ROC) among the eligible models (see “Predictive modeling”). Using the chosen thresholds closest to 95% specificity in our data, we also computed sensitivity/recall or true positive rate (TPR), precision or positive predictive value, negative predictive value (NPV), and specificity or true negative rate (TNR). Eventually, we evaluated the model explainability and interpretability. The former was inferred by the Shapley additive explanation (SHAP) beeswarm plot, while the latter was inferred by regression analysis using the predicted outcome as the dependent variable. The concordance was visually assessed between the feature impact on model output based on the SHAP beeswarm plot and the causal effect size based on the multivariate regression.

In addition, we conducted an error analysis to understand the best model. To select examples that were representative for the error analysis, we randomly selected a sample for each cluster among true and false positives and negatives. The samples were clustered by *k*-mean algorithm. We chose *k*-value *post hoc* based on principal component analysis with original and reduced dimensional ontologies using hierarchical clustering.

The source codes were publicly shared (https://github.com/herdiantrisufriyana/fatigue_hba1c) for reproducing our data analysis. All the analyses were conducted using R version 4.4.1, except for model training and explainability which used Python version 3.12.4. We provided docker to replicate our programming environment.

## Results

### Sample characteristics

We have enrolled 281 participants in this study (Table 1). There were 40.56% (*n*=114/281) of participants who reported fatigue. These participants included individuals with type 2 diabetes for 5.30 (95% CI 1.50, 9.10) years, who received oral hypoglycemic agent (OHA) last 3 months. They were 52.46 (95% CI 41.45, 63.48) years old with minimum and maximum HbA1c of 5.9 and 10.1, respectively. Our samples who reported or did not report fatigue included all the categories of sex, educational status, marital status, comorbidity, and cigarette smoking status.

**Table 1.** Sample characteristics.

| Variable |  | Fatigue |  |
| --- | --- | --- | --- |
|  |  | no (n=167) | yes (n=114) |
| HbA1c (%) | average (SD) | 6.65 (0.39) | 7.68 (0.88) |
|  | 5.7 to <6.5 % (n) | 33 (55) | 8 (9) |
|  | 6.5+ % (n) | 67 (112) | 92 (105) |
| Age (years) | average (SD) | 52.82 (5.14) | 52.46 (5.62) |
| Sex | male % (n) | 34 (56) | 30 (34) |
|  | female % (n) | 66 (111) | 70 (80) |
| Educational status | no education % (n) | 4 (7) | 4 (5) |
|  | primary school % (n) | 27 (45) | 37 (42) |
|  | junior secondary school % (n) | 28 (47) | 30 (34) |
|  | senior secondary school % (n) | 23 (38) | 15 (17) |
|  | college graduate % (n) | 18 (30) | 14 (16) |
| DM duration (years) | average (SD) | 5.43 (2.07) | 5.30 (1.94) |
| Marital status | unmarried % (n) | 5 (8) | 4 (4) |
|  | married % (n) | 71 (119) | 73 (83) |
|  | widowed/divorced % (n) | 24 (40) | 24 (27) |
| Comorbidity | no % (n) | 72 (121) | 10 (11) |
|  | yes % (n) | 28 (46) | 90 (103) |
| OHA + insulin | OHA % (n) | 86 (143) | 62 (71) |
|  | OHA + insulin % (n) | 14 (24) | 38 (43) |
| Cigarette smoking | no % (n) | 90 (150) | 87 (99) |
|  | yes % (n) | 10 (17) | 13 (15) |

DM, diabetes mellitus; OHA, oral hypoglycemic agent; SD, standard deviation.

### Structural causal modeling

We identified 11 relationships with statistical significances between a pair of variables (Figure S1), excluding missingness and redundant variables (e.g., HbA1c and its transformation). The causal directions were identified based on the LLM responses (Table S3). Therefore, we could construct a causal diagram (Figure 1) to determine covariates in a multivariate regression analysis for the variables that were correlated to fatigue with causal directions from each variable to fatigue.

**Figure 1.**
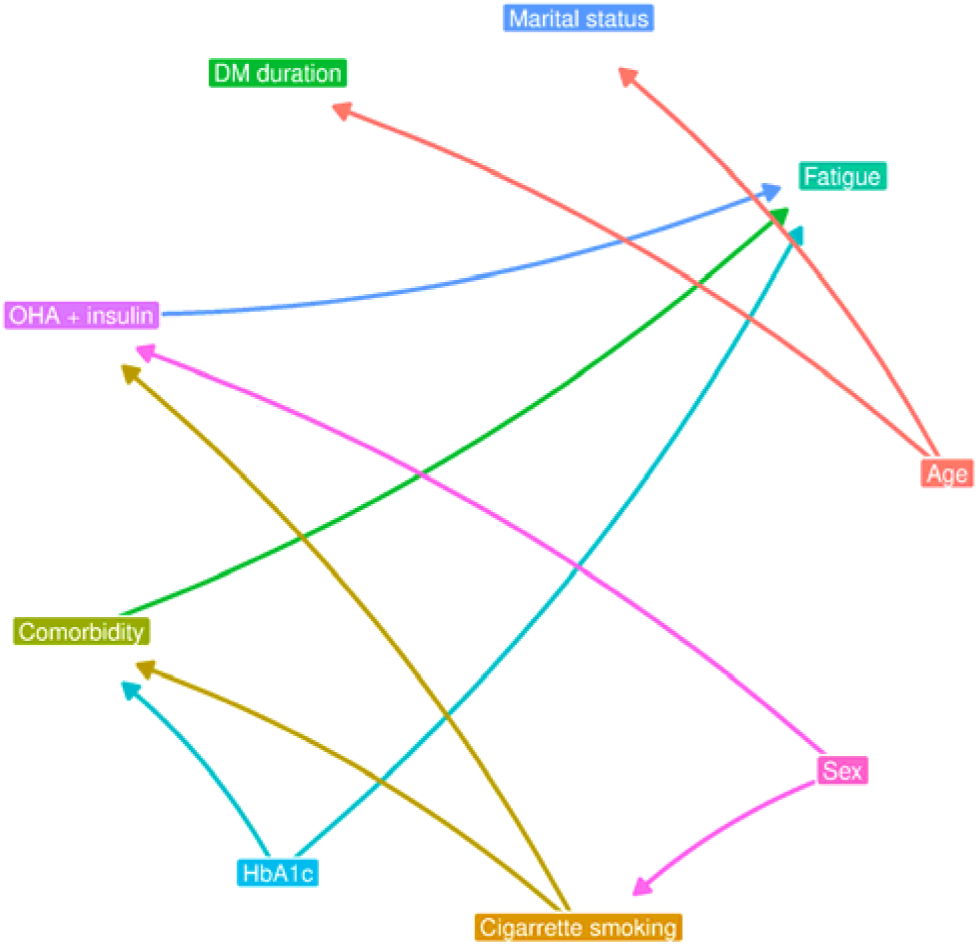
Causal diagram based on correlation matrix and LLM responses. DM, diabetes mellitus; LLM, large language model; OHA, oral hypoglycemic agent.

In a multivariate regression analysis (Table S4), the covariates for each variable included only confounders and mediators if any. Individuals with poor HbA1c levels, i.e., ≥6.5%, was 6.1 times (95% CI 2.5, 14.4; *p*-value <0.001) more likely to report fatigue, compared to those with HbA1c 5.7% to < 6.5%, after adjustment for comorbidity as mediator. Meanwhile, individuals with comorbidity last 3 months was 25.2 times (95% CI 12.1, 52.4; *p*-value <0.001) more likely to report fatigue, compared to those without comorbidity, after adjustment for poor HbA1c levels. If individuals required insulin last 3 months in addition to OHA, they were 3.6 times (95% CI 2.0, 6.4; *p*-value <0.001) more likely to report fatigue. Therefore, we selected HbA1c level, comorbidity, and DM treatment as the candidate predictors for predictive modeling.

### Predictive modeling

We trained three prediction models using the three predictors and three ML algorithms that fulfilled sufficient sample size estimation (Figure S2): (1) KNN; (2) DT; and (3) RF. These models were also well-calibrated (Figures 2A-C) and had positive net benefits using thresholds closest to 95% specificity in our data (Figure 2D). Using these thresholds, the ROC curve (Figure 2E) showed that the three had similar sensitivities. However, the interval estimates (Table 2) indicated that KNN resulted in the most sensitive model (TNR=95.1%, 95% CI 94.5, 95.7) while the most precise model was resulted from either DT (PPV=91.4%, 95% CI 90.5, 92.3) or RF (PPV=90.6%, 95% CI 89.6, 91.6).

**Figure 2.**
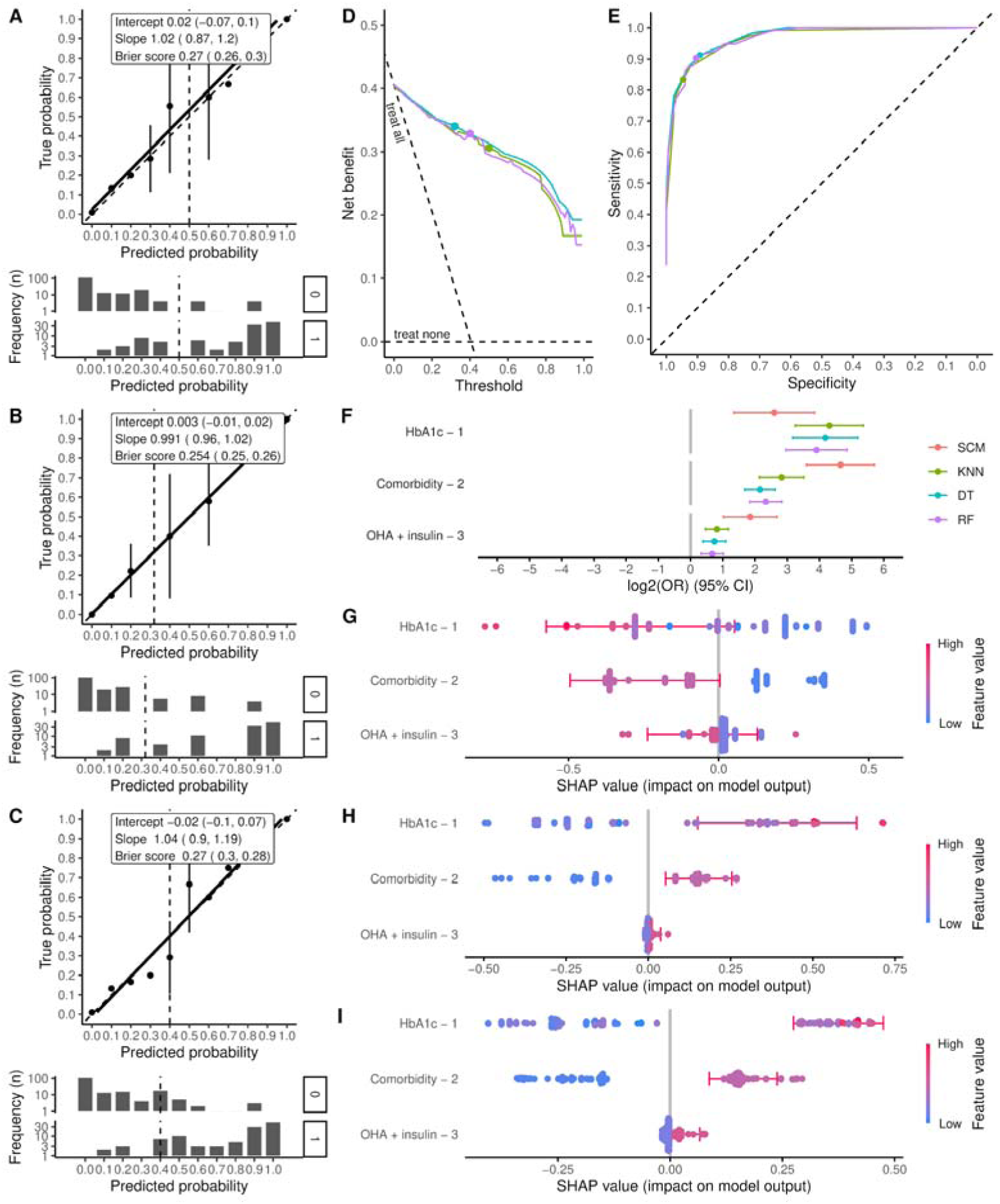
Model evaluation for calibration (A-C), clinical utility (D), discrimination ability (E), and explainability and interpretability (F-J). Calibration and SHAP beeswarm plots are shown for KNN (A and G), DT (B and H), and RF (C and I). In calibration plots (A-C), the values 0 and 1 refer to fatigue values of no and yes, respectively. Color legends indicating the models are shared among decision curve (D), ROC curve (E), and regression analysis of the predictors using the predicted outcome as the dependent variable (F). Points in decision and ROC curves (D, E) correspond to the chosen threshold closest to 95% specificity in our data. CI, confidence interval; DM, diabetes mellitus; DT, decision tree; KNN, k-nearest neighbor; OHA, oral hypoglycemic agent; OR, odds ration; RF, random forest; ROC, receiver operating characteristics; SCM, structural causal model; SHAP, Shapley additive explanation.

**Table 2.** Predictive performance.

| Model | AUC-ROC | Th. | Sensitivity/recall<br>or TPR (%) | Precision or<br>PPV (%) | NPV (%) | Specificity or<br>TNR (%) |
| --- | --- | --- | --- | --- | --- | --- |
| KNN | 0.964 (0.959, 0.968) | 0.50 | 95.1 (94.5, 95.7) | 82.2 (80.8, 83.6) | 91.9 (90.9, 92.9) | 88.8 (87.9, 89.7) |
| DT | 0.971 (0.967, 0.974) | 0.32 | 89.7 (88.9, 90.4) | 91.4 (90.5, 92.3) | 85.7 (84.8, 86.6) | 93.9 (93.2, 94.6) |
| RF | 0.966 (0.962, 0.969) | 0.40 | 90.7 (90.0, 91.4) | 90.6 (89.6, 91.6) | 86.9 (86.0, 87.7) | 93.4 (92.7, 94.2) |

AUC-ROC, area under the curve - receiver operating characteristics; DT, decision tree; KNN, *k*-nearest neighbor; Th., the chosen threshold closest to 95% specificity in our data; NPV, negative predictive value; PPV, positive predictive value; RF, random forest; TNR, true negative rate; TPR, true positive rate.

Nevertheless, KNN had poor concordance between the feature impact on model output (Figure 2G) and the causal effect size in SCM (Figure 2F), unlike DT (Figure 2H) and RF (Figure 2I). Eventually, we chose RF as the best model *post hoc*, although the best model was resulted from either DT or RF. It is because the predicted probabilities were evenly distributed using RF. The AUC-ROC of the best model was 0.966 (95% CI 0.962, 0.969).

For an error analysis, we identified sample clusters from which the representative examples were selected among true and false positives and negatives. Most of variance (80%) in our data was explained by a principal that was almost completely determined by age (Figure S3). We used this variable to identify the clusters (Figure S4): (1) age ≤53 years old; and (2) age >53 years old. In the latter cluster (Figure S5), two examples were similarly predicted as negatives with similar patterns of the predictor impacts on the model output, but an example was a false negative. Older participants might require variables beyond those in this study to reduce the number of false negatives and improve detection rate (i.e., sensitivity).

### Model usage

We have provided the workflow, web application (https://predme.app/fatigue_hba1c), and nomogram to use our model. Our model is used for an individual with type 2 diabetes who reports fatigue. The individual also receives oral hypoglycemic agent last 3 months without or with insulin/comorbidity (Figure 3A). This information and HbA1c level must be sourced from medical records. Since our model detected ∼90% of participants who reported fatigue, it may not be detected in ∼10% individuals. In this case, our model cannot be used to identify whether current fatigue is related to poor HbA1c. A user is suggested to seek other causes of fatigue beyond HbA1c and comorbidity.

**Figure 3.**
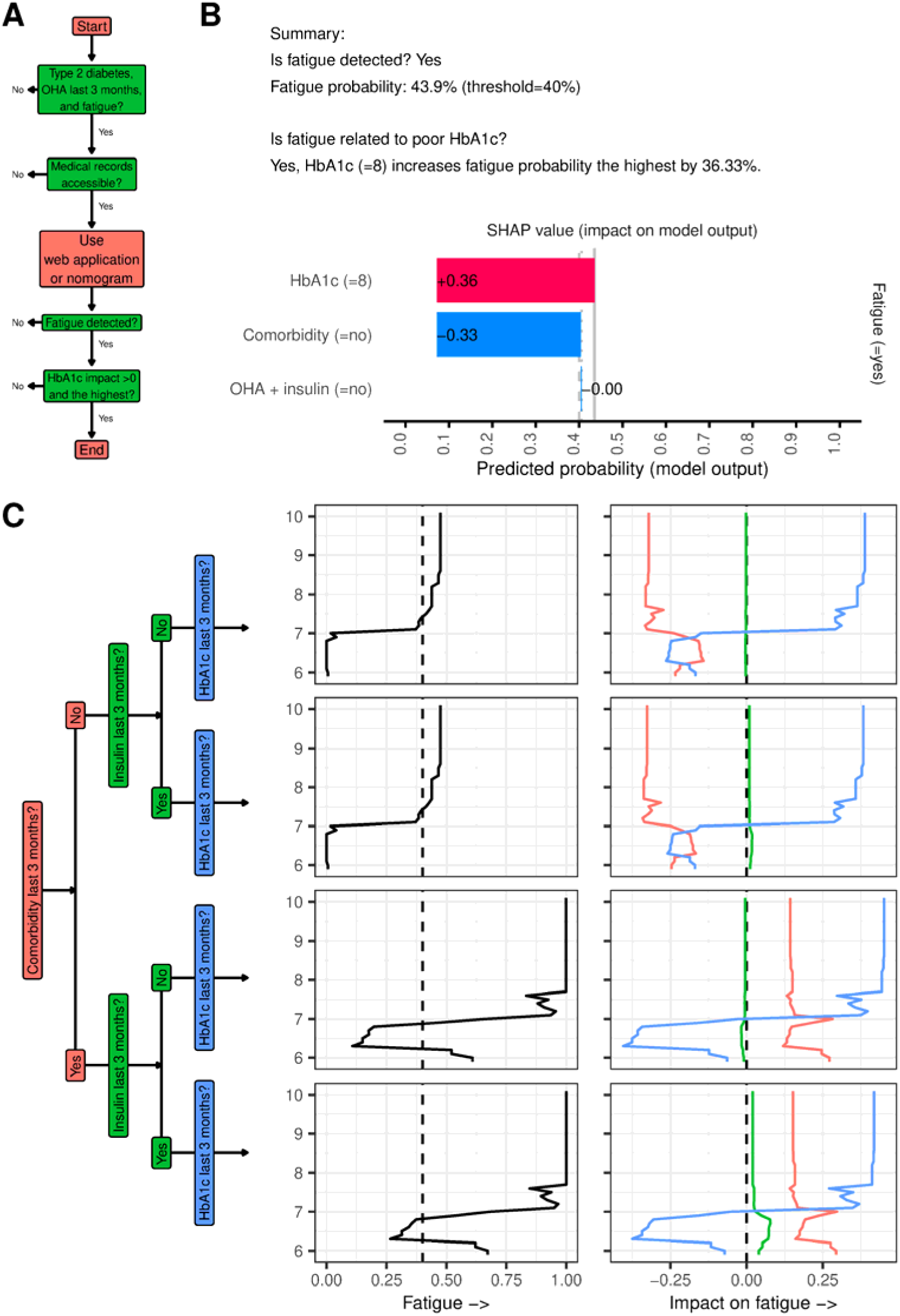
Workflow (A), web application (B), and nomogram (C) to use our model. Workflow (A) is red and green respectively for start/process/end and decision. Web application (B) is shown for an example case. In nomogram (C), the line colors for the impact on fatigue correspond to the box colors in the decision tree, indicating the variables. OHA, oral hypoglycemic agent.

As an example, the interface of prediction results of the web application is shown for an eligible individual whose fatigue is detected (Figure 3B). In this example, fatigue is related to poor HbA1c because its impact is >0 and the highest among the predictors, as indicated in the workflow (Figure 3A). Alternatively, a user can use nomogram in a setting where internet access is not reliable (Figure 3C). In addition, if HbA1c level is unknown, a user can use this nomogram to estimate the level. The example case may have HbA1c level >7% based on this nomogram. A clinician may order HbA1c check and identify whether fatigue is related to the HbA1c level.

## Discussion

### Finding summary

Individuals with type 2 diabetes receiving OHA last 3 months were more likely to report fatigue when they had poor HbA1c (adjusted OR [aOR]=6.1, 95% CI 2.5, 14.4), comorbidity (aOR=25.2, 95% CI 12.1, 52.4), or a need for insulin treatment (aOR=3.6, 95% CI 2.0, 6.4). We have developed a web application and nomogram using the three variables to identify whether HbA1c level contributes to fatigue for each individual case among 95.1% (95% CI 94.5, 95.7) of those who reported fatigue. In addition, a nomogram was also developed and it can be used to estimate the HbA1c level.

### Research and clinical implications

This study demonstrated a feasibility to engineer SCM and ML model explainability for identifying a symptom-related cause at individual level. While there was no previous study that applies covariance-based causal analysis to approach specific causality, such effort is similar to stratified medicine for treatment-response prediction, a term for targeting a treatment based an absolute risk for each individual case [30]. However, stratified medicine applies statistical inference which does not incorporate domain knowledge systematically for inferring the causal structure as SCM does [31]. Data-driven approach alone may encounter more type 1 errors, because the predictors include common effects or confounders/mediators that are affected by the common effects [25]. Moreover, SCM with ML model explainability allows more training algorithms, not limited to general additive modeling such as linear/logistic regression, for optimizing the model accuracy. A new algorithm by future research in this field is also expected to improve the concordance between the feature impact on model output and the effect size based on the SCM.

For clinical application, identifying fatigue-related poor HbA1c demonstrated an example of data-driven, decision-making support that assists clinicians in prioritizing the follow-up. It is important particularly in healthcare funded by a capitation model which accrued higher costs among individuals with type 2 diabetes, compared to those of fee-for-service model [32]. Specifically, a prioritization is important between modifying the treatment for improving HbA1c and addressing the comorbidity. Treatment costs contributed to the largest difference of costs between individuals with and without type 2 diabetes [33, 34]. The major comorbidity of type 2 diabetes, i.e., cardiovascular diseases, contributed to higher costs even larger [34, 35]. Therefore, identifying fatigue-related poor HbA1c is not only helpful to address this symptom and its impact on DSM [5], and subsequently avoid the complications of poor HbA1c [6], but also reduce the costs.

### Strength and limitations

This study utilized two objective methods for SCM to infer the causal direction using knowledge base via LLM and to estimate the effect using data via multivariate regression analysis. The former method enables an internet-size coverage for inferring causal direction which minimizes type 1 errors due to inclusion of predictors from common effects or confounders/mediators that are affected by the common effects. Our web application and nomogram provide a quantity of a predictor impact on the model output; thus, a clinician can prioritize a follow-up to address fatigue in type diabetes. A clinician is also informed about the estimated HbA1c using our nomogram to make a decision in ordering HbA1c check.

We also considered several limitations of this study. First, our data did not include several factors of fatigue in type 2 diabetes, e.g., depression [1, 8] and sleep quality [7]. These factors might contribute the false negatives of the two examples, as shown in our error analysis. However, our model still informs a clinician to identify other causes, including depression and sleep, particularly in a case with fatigue undetected by our model. Second, our model accuracy is not externally validated, yet. Nevertheless, we provided an open-access web application and nomogram for other researchers to validate our model in their populations. Third, SCM in this study might not cover all the confounders between each predictor and fatigue. While it is impossible to exhaustively include the confounders, we have provided the cause identifier of fatigue to be examined for its validity in future RCTs.

### Conclusion and future studies

We have developed a fatigue prediction model that could be used to determine whether HbA1c level contributes to fatigue for each individual case. Our model is accessible via web application and nomogram, and detected most individuals with type diabetes who reported fatigue. Information of the estimated HbA1c was also provided by our nomogram for considering whether HbA1c check is needed.

Future studies are expected before applying our web application and nomogram in real-world settings. First, an external validation is needed to validate our model accuracy. Second, the validity of the cause identifier of fatigue must be examined by RCT. Third, an impact study is also needed to assess the benefits of using our web application and nomogram in improving the patient outcomes.

## Supporting information

TRIPOD+AI Checklist

## Acknowledgments

This study was funded by: (1) the Postdoctoral Accompanies Research Project from the National Science and Technology Council (NSTC) of Taiwan (grant nos.: NSTC111-2811-E-038-003-MY2 and NSTC113-2811-E-A49A-003) to HS; (2) the Exchange Faculty Travel Grant under the Ratchadaphiseksomphot Fund, Chulalongkorn University (The Exchange Faculty Travel Grant; Grant No. CTG167005) to DSR; and (3) the National Science and Technology Council in Taiwan (grant no. NSTC113-2221-E-A49-193-MY3), the Ministry of Science and Technology (MOST) of Taiwan (grant nos.: MOST110-2628-E-038-001 and MOST111-2628-E-038-001-MY2), the University System of Taipei Joint Research Program (grant no.: USTP-NTOU-TMU-112-04), and the Higher Education Sprout Project from the Ministry of Education (MOE) of Taiwan (grant no.: DP2-111-21121-01-A-05 and DP2-TMU-112-A-13) to ECYS. These funding bodies had no role in the study design; in the collection, analysis, and interpretation of data; in the writing of the report; or in the decision to submit the article for publication.

## CRediT authorship

**HS:** Conceptualization, Methodology, Software, Validation, Formal analysis, Writing – original draft, Visualization, Funding acquisition. **DSR:** Conceptualization, Methodology, Validation, Investigation, Data curation, Writing – original draft, Project administration, Funding acquisition. **RK:** Conceptualization, Investigation, Resources, Data curation, Writing – review & editing. **SA:** Investigation, Data curation, Writing – review & editing, Project administration. **EE:** Investigation, Data curation, Writing – review & editing, Project administration. **EPO:** Investigation, Data curation, Writing – review & editing, Project administration. **HYC:** Conceptualization, Methodology, Writing – review & editing, Supervision, Funding acquisition. **ECYS:** Conceptualization, Methodology, Resources, Writing – review & editing, Supervision, Funding acquisition. All authors have read and approved the manuscript and agreed to be accountable for all aspects of the work in ensuring that questions related to the accuracy or integrity of any part of the work are appropriately investigated and resolved.

## Data availability

Data are available publicly (https://doi.org/10.5281/zenodo.14848166). We also publicly shared the source codes for data analysis (https://github.com/herdiantrisufriyana/fatigue_hba1c). An open-access web application (https://predme.app/fatigue_hba1c) is provided for using our model.

## References

1. Romadlon, D.S., et al., Prevalence and risk factors of fatigue in type 1 and type 2 diabetes: A systematic review and meta-analysis. J Nurs Scholarsh, 2022. 54(5): p. 546–553.

2. Romadlon, D.S., et al., Fatigue following type 2 diabetes: Psychometric testing of the Indonesian version of the multidimensional fatigue Inventory-20 and unmet fatigue-related needs. PLoS One, 2022. 17(11): p. e0278165.

3. Fritschi, C. and L. Quinn, Fatigue in patients with diabetes: a review. J Psychosom Res, 2010. 69(1): p. 33–41.

4. Singh, R., et al., Fatigue in Type 2 Diabetes: Impact on Quality of Life and Predictors. PLoS One, 2016. 11(11): p. e0165652.

5. Kuo, H.J., et al., Impact of Fatigue and Its Influencing Factors on Diabetes Self-Management in Adults With Type 2 Diabetes: A Structural Equation Modeling Analysis. Sci Diabetes Self Manag Care, 2023. 49(6): p. 438–448.

6. Romadlon, D.S., et al., Comparative Effects of Diabetes Self-Management Programs on Type 2 Diabetes Clinical Outcomes: A Systematic Review and Network Meta-Analysis. Diabetes Metab Res Rev, 2024. 40(6): p. e3840.

7. Bi, Y., et al., Contributing factors of fatigue in patients with type 2 diabetes: A systematic review. Psychoneuroendocrinology, 2021. 130: p. 105280.

8. Kuo, H.J., Y.C. Huang, and A.A. García, An integrative review of fatigue in adults with type 2 diabetes mellitus: Implications for self-management and quality of life. J Clin Nurs, 2022. 31(11-12): p. 1409–1427.

9. Ma, Y., et al., Prevalence and risk factors of cancer-related fatigue: A systematic review and meta-analysis. Int J Nurs Stud, 2020. 111: p. 103707.

10. Aali, G., et al., Post-stroke fatigue: a scoping review. F1000Res, 2020. 9: p. 242.

11. Oliva Ramirez, A., et al., Prevalence and burden of multiple sclerosis-related fatigue: a systematic literature review. BMC Neurol, 2021. 21(1): p. 468.

12. Pavlovic, N.V., et al., Fatigue in Persons With Heart Failure: A Systematic Literature Review and Meta-Synthesis Using the Biopsychosocial Model of Health. J Card Fail, 2022. 28(2): p. 283–315.

13. Siciliano, M., et al., Fatigue in Parkinson’s disease: A systematic review and meta-analysis. Mov Disord, 2018. 33(11): p. 1712–1723.

14. Hamidovic, A. and H. de Wit, Sleep deprivation increases cigarette smoking. Pharmacol Biochem Behav, 2009. 93(3): p. 263–9.

15. Corwin, E.J., L.C. Klein, and K. Rickelman, Predictors of fatigue in healthy young adults: moderating effects of cigarette smoking and gender. Biol Res Nurs, 2002. 3(4): p. 222–33.

16. Kahraman, T., et al., Associations between smoking and walking, fatigue, depression, and health-related quality of life in persons with multiple sclerosis. Acta Neurol Belg, 2021. 121(5): p. 1199–1206.

17. Halpern, J.Y., Actual causality. 2016: MiT Press.

18. Hellner, J., Causality and causation in law. Scandinavian studies in law, 2000. 40: p. 111–134.

19. Peters, J., D. Janzing, and B. Schölkopf, Elements of causal inference: foundations and learning algorithms. 2017: The MIT Press.

20. Imbens, G.W. and D.B. Rubin, Causal inference in statistics, social, and biomedical sciences. 2015: Cambridge university press.

21. Hernán, M.A. and J.M. Robins, Causal inference. 2010, CRC Boca Raton, FL.

22. Geffner, H., R. Dechter, and J.Y. Halpern, Probabilistic and Causal Inference: The Works of Judea Pearl. 2022: ACM.

23. Hernán, M.A., Methods of Public Health Research - Strengthening Causal Inference from Observational Data. N Engl J Med, 2021. 385(15): p. 1345–1348.

24. Zabor, E.C., A.M. Kaizer, and B.P. Hobbs, Randomized Controlled Trials. Chest, 2020. 158(1s): p. S79–s87.

25. Pearl, J. and D. Mackenzie, The book of why: the new science of cause and effect. 2018: Basic books.

26. Ali, S., et al., Explainable Artificial Intelligence (XAI): What we know and what is left to attain Trustworthy Artificial Intelligence. Information fusion, 2023. 99: p. 101805.

27. Kıcıman, E., et al., Causal reasoning and large language models: Opening a new frontier for causality. arXiv preprint arXiv:2305.00050, 2023.

28. van der Ploeg, T., P.C. Austin, and E.W. Steyerberg, Modern modelling techniques are data hungry: a simulation study for predicting dichotomous endpoints. BMC Med Res Methodol, 2014. 14: p. 137.

29. Sufriyana, H. and E.C.-Y. Su, rmlnomogram: An R package to construct an explainable nomogram for any machine learning algorithms. arXiv preprint arXiv:2501.05772, 2025.

30. Hingorani, A.D., et al., Prognosis research strategy (PROGRESS) 4: stratified medicine research. Bmj, 2013. 346: p. e5793.

31. Riley, R.D., et al., Prognosis Research Strategy (PROGRESS) 2: prognostic factor research. PLoS Med, 2013. 10(2): p. e1001380.

32. Naidoo, L.A., et al., Is the Risk Really Shared? A Retrospective Analysis of Healthcare Costs of Patients With Type 2 Diabetes Mellitus on a Capitation Model. Value Health Reg Issues, 2022. 28: p. 29-37.

33. Jacobs, E., et al., Healthcare costs of Type 2 diabetes in Germany. Diabet Med, 2017. 34(6): p. 855–861.

34. Wu, J.H., et al., Healthcare Costs Associated with Complications in Patients with Type 2 Diabetes among 1.85 Million Adults in Beijing, China. Int J Environ Res Public Health, 2021. 18(7).

35. Einarson, T.R., et al., Economic Burden of Cardiovascular Disease in Type 2 Diabetes: A Systematic Review. Value Health, 2018. 21(7): p. 881–890.

