## Supplemental Materials for "Large language model-assisted causal machine learning for identifying fatigue-related poor glycated hemoglobin in type 2 diabetes"

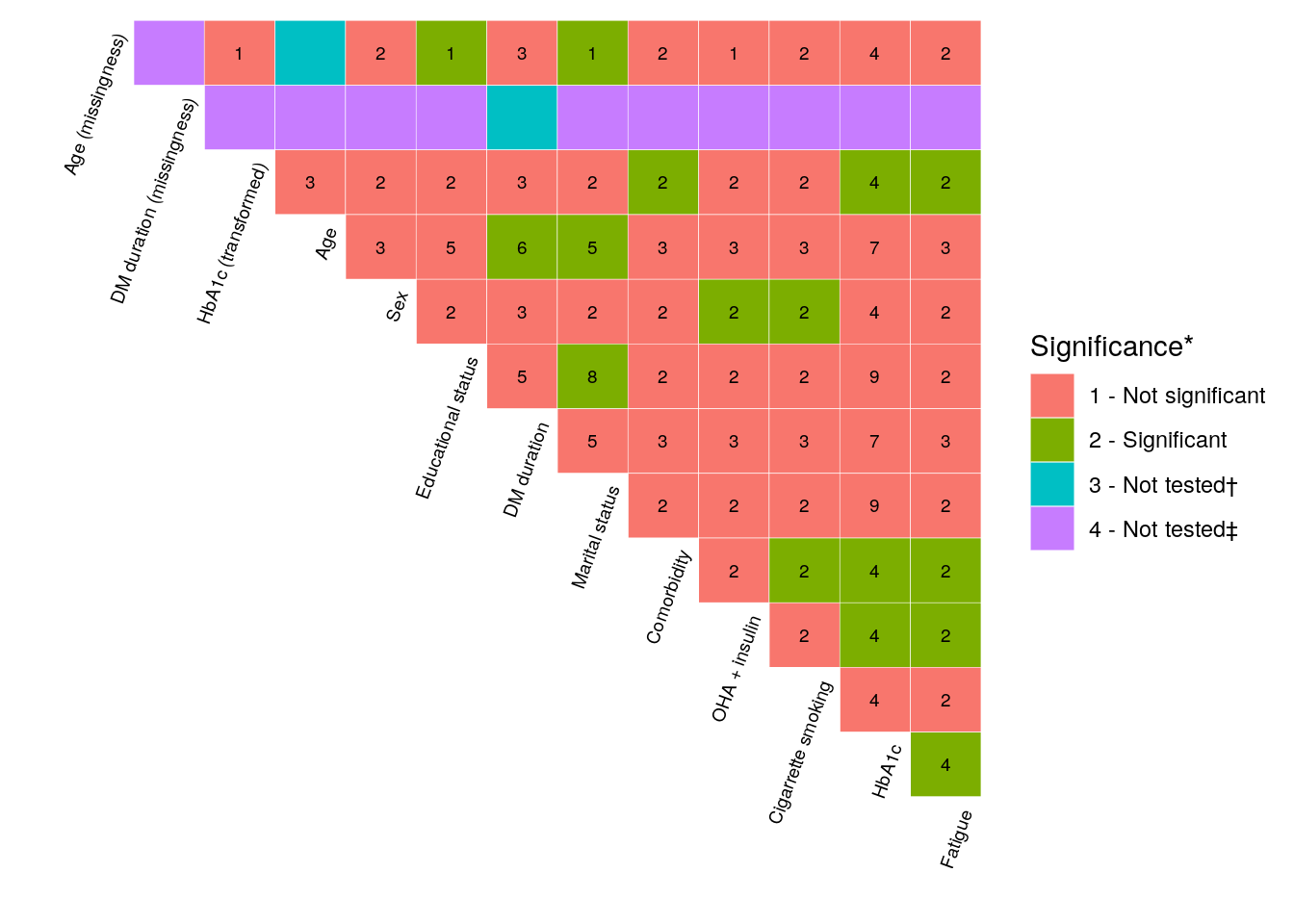

Figure S1. Correlation matrix. The numbers indicate the statistical tests: (1) Bayesian binomial logistic regression analysis; (2) Fisher's exact test; (3) Welch's *t*-test; (4) Mann-Whitney *U* test (Wilcoxon rank-sum test); (5) ANOVA; (6) Pearson's correlation test; (7) Spearman's correlation test; (8) Bayesian multinomial logistic regression analysis; and (9) Kruskal-Wallis test. *, based on *p*-value (frequentist) or 95% CI (Bayesian); †, a variable and its missingness; ‡, at least 1 variable was a categorical variable with a category of 1 value. ANOVA, analysis of variance.

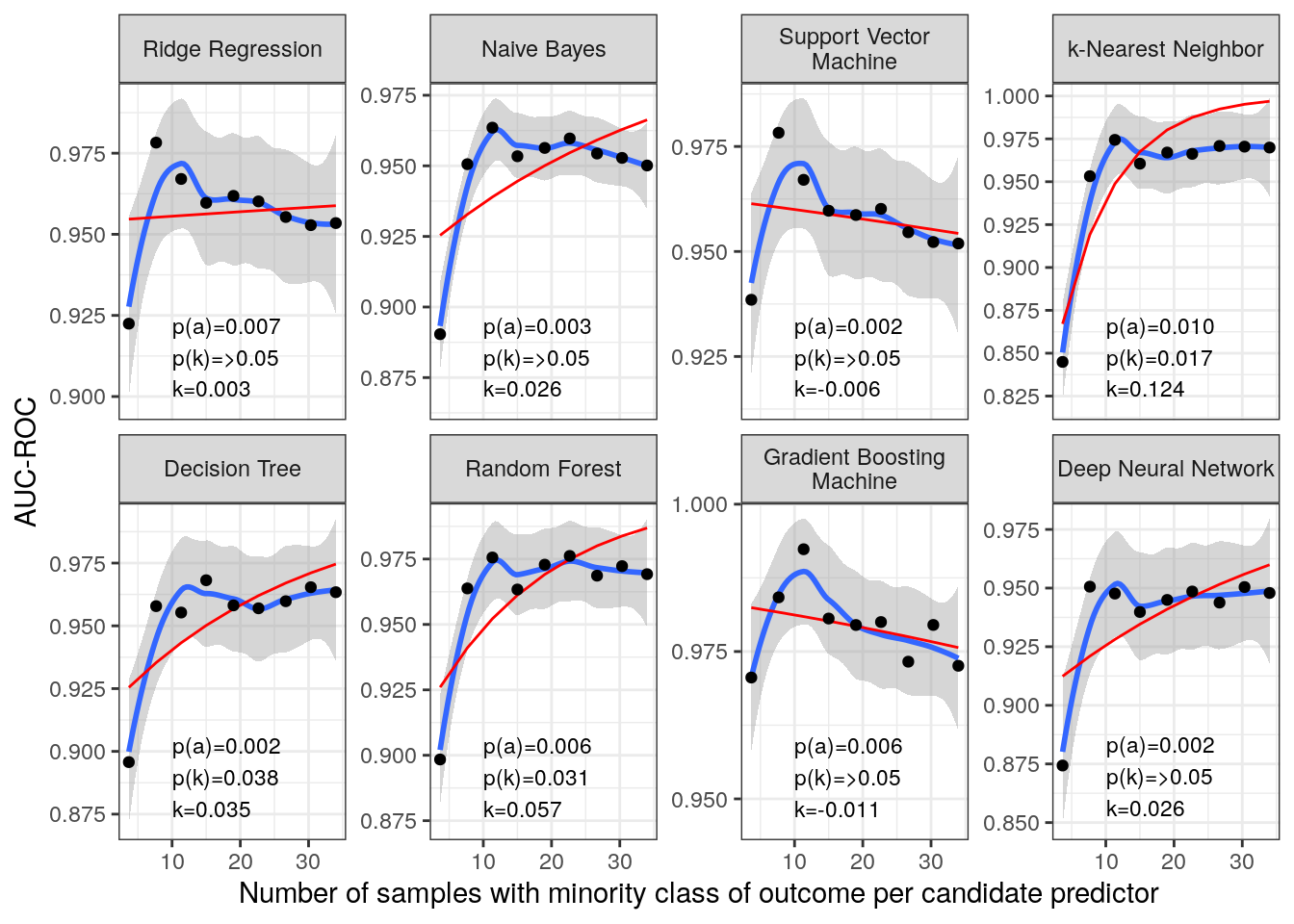

Figure S2. Sample size estimation. Blue line indicates the LOESS. Red line indicates the modified exponential decay fitting. *p*(*a*) is the *p*-value of a where *a* is a parameter represents the factor by which the exponential term was scaled in the model and a higher value generally means a quicker initial increase. *p*(*k*) is the *p*-value of *k* where *k* is the rate of decay or growth in the exponential term and a positive *k* implies a decay (as *x* increases, the impact of increasing *y* diminishes) while a negative *k* suggests an erroneous model fit in this context because an increase in *x* should not lead to a decrease in *y*. AUC-ROC, the area under curve of receiver operating characteristics; LOESS, locally estimated scatterplot smoothing.

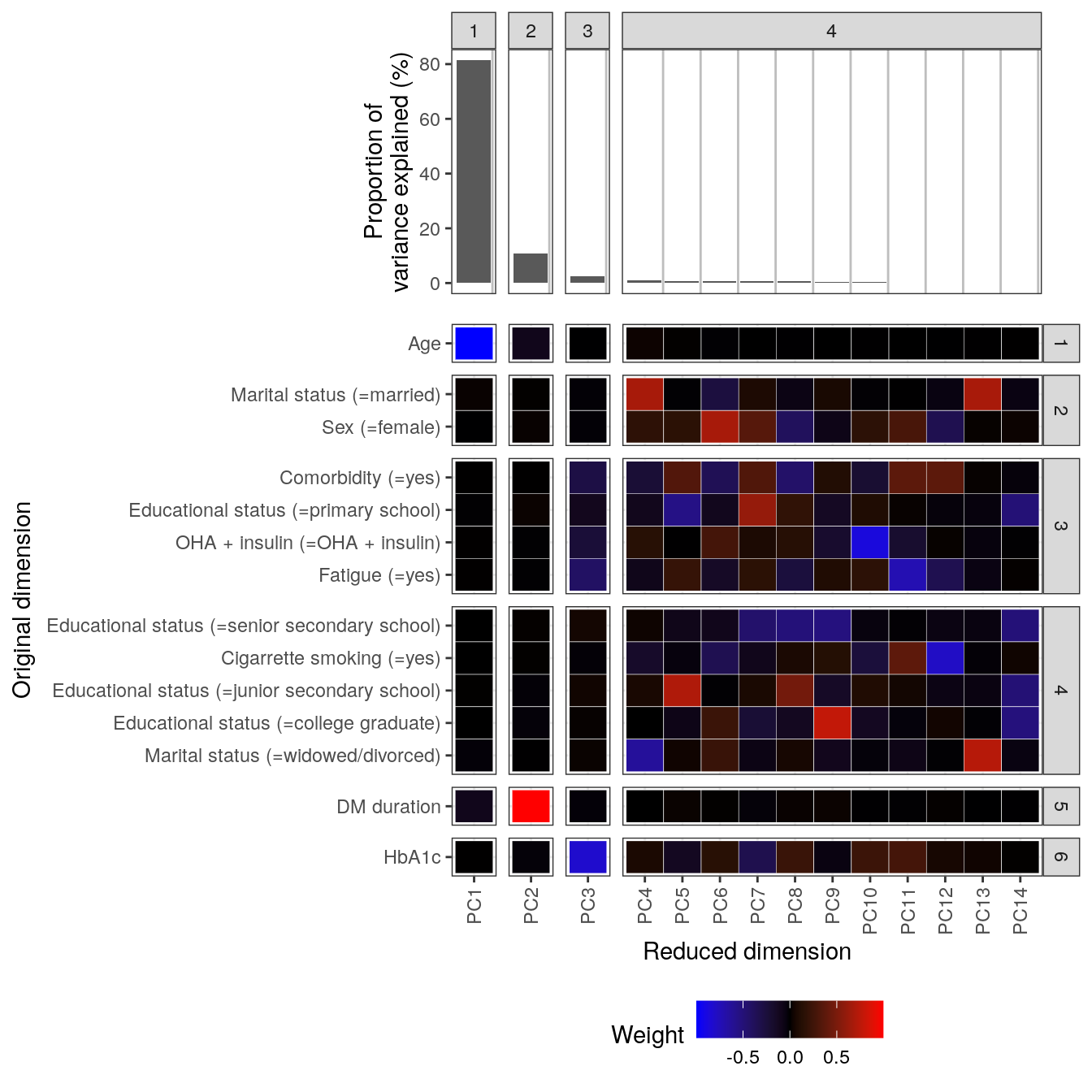

Figure S3. Principal component analysis with original and reduced dimensional ontologies using hierarchical clustering. Ontologies were derived by cutting hierarchical cluster tree such that the resulting groups demonstrated meaningful pattern. It was determined based on visual assessment of the weights and semantic assessment of the original dimensions. The former resulted in six ontologies of the reduced dimensions, while the latter resulted in four ontologies of the original dimensions. DM, diabetes mellitus; OHA, oral hypoglycemic agent.

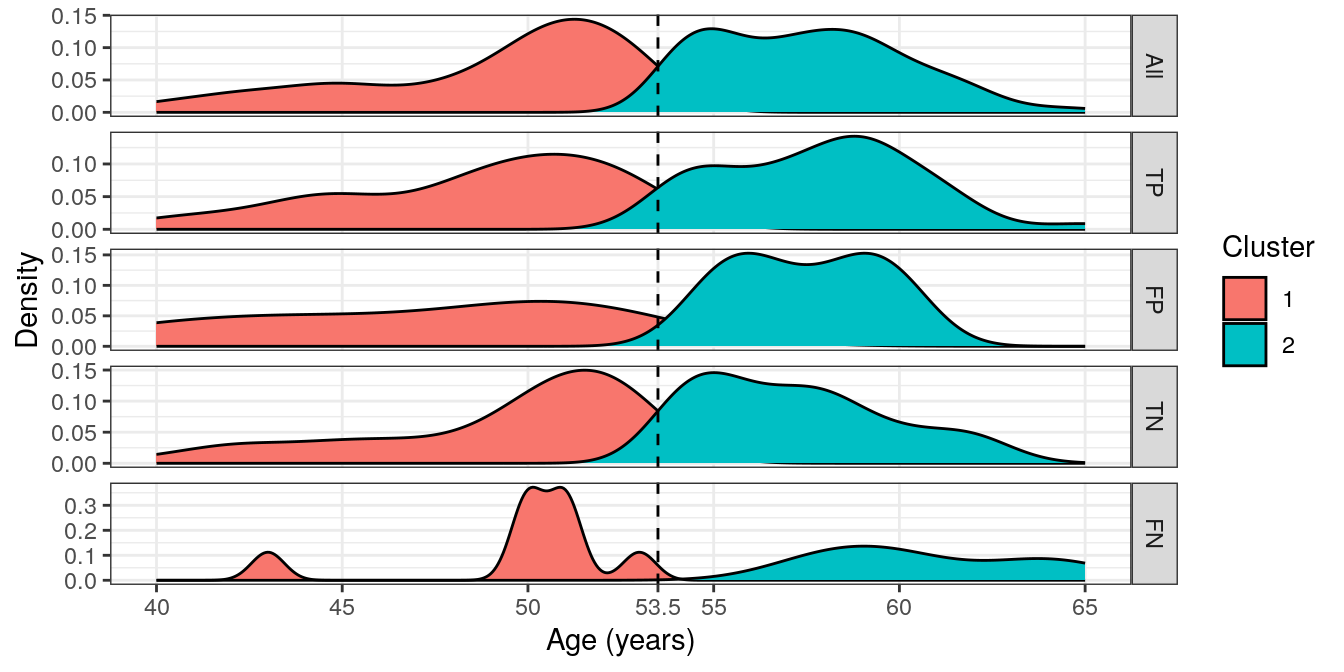

Figure S4. Sample clustering by *k*-mean. We selected *k*=2 *post hoc* because it provided samples for each cluster among true and false positives and negatives, completely. Age was selected for clustering the samples because it almost singlehandedly determined a principal component that had the highest proportion of the variance explained in our data. FN, false negative; FP, false positive; TN, true negative; TP, true positive.

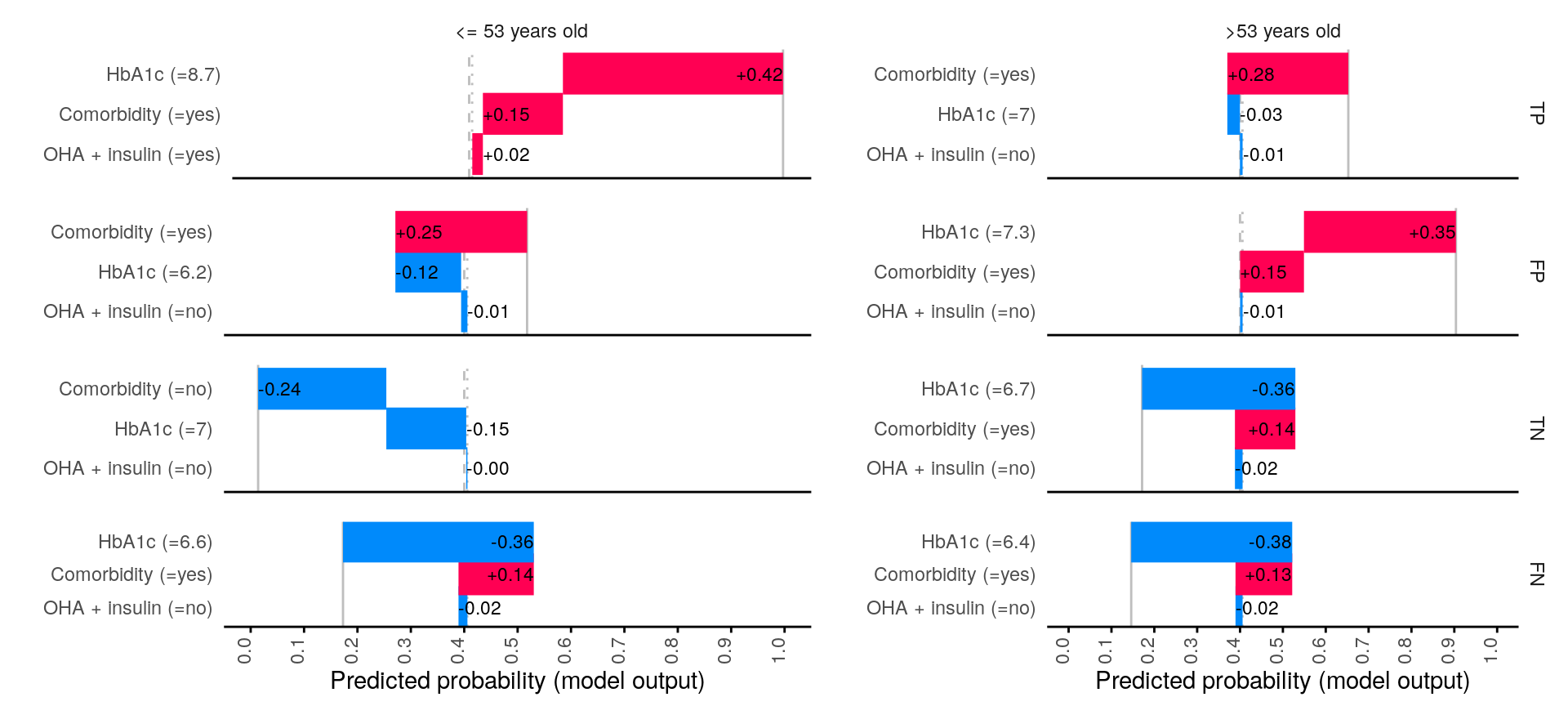

Figure S5. Error analysis on the selected examples by SHAP waterfall plots. SHAP, Shapley additive explanation. OHA, oral hypoglycemic agent.

Table S1. Reasoning for the algorithm of choices.

| Algorithm | Reason | | | | | |
| --- | --- | --- | --- | --- | --- | --- |
|  | Linear relationship | Optimal for categorical predictors | High dimensionality | Interpretable | Sensitive to hyperparameter | High noise tolerance |
| Ridge regression | Yes | Yes | Yes | Yes | No | No |
| Naïve Bayes | No | Yes | Yes | Yes | No | No |
| Support vector machine with linear kernel | Yes | No | Yes | No | Yes | No |
| *k*-nearest neighbor | No | Yes | No | No | No | No |
| Decision tree | No | Yes | No | Yes | Yes | No |
| Random forest | No | Yes | Yes | No | No | Yes |
| Gradient boosting machine | No | Yes | Yes | No | Yes | No |
| Deep neural network | No | Yes | Yes | No | Yes | Yes |

Table S2. Algorithm hyperparameters.

| Algorithm | Hyperparameters | |
| --- | --- | --- |
|  | Function/  Argument=fixed value/  Argument: possible values | Note |
| Ridge regression | LogisticRegression |  |
|  | penalty='l2' |  |
|  | solver='lbfgs' |  |
|  | max_iter=1000 |  |
|  | 'C': np.logspace(-4, 4, 10) | Regularization strength |
| Naïve Bayes | GaussianNB | Gaussian Naive Bayes |
|  | N/A | Naive Bayes typically doesn't require hyperparameter tuning |
| Support vector machine with linear kernel | SVC |  |
|  | kernel='linear' |  |
|  | probability=True | Enable probability estimates |
|  | 'C': np.logspace(-4, 4, 10) | Penalty parameter C of the error term |
| *k*-nearest neighbor | KNeighborsClassifier |  |
|  | 'n_neighbors': [3, 5, 7, 9] | Number of neighbors |
|  | 'weights': ['uniform', 'distance'] | Weight function used in prediction |
|  | 'metric': ['euclidean', 'manhattan'] | Distance metric for tree construction |
| Decision tree | DecisionTreeClassifier |  |
|  | min_samples_split=20 |  |
|  | 'max_depth': [3, 5, 10] | Maximum depth of trees |
| Random forest | RandomForestClassifier |  |
|  | min_samples_split=20 |  |
|  | max_features=None |  |
|  | 'n_estimators': [100, 200, 300] | Number of trees |
|  | 'max_depth': [3, 5, 10] | Maximum depth of trees |
| Gradient boosting machine | GradientBoostingClassifier |  |
|  | learning_rate=0.01 |  |
|  | 'n_estimators': [100, 200, 300] | Number of boosting stages |
|  | 'max_depth': [3, 5, 10] | Max depth of regression estimators |
| Deep neural network | MLPClassifier |  |
|  | max_iter=5000 |  |
|  | hidden_layer_sizes=(10, 10) |  |
|  | activation='relu' |  |
|  | solver='sgd' |  |
|  | learning_rate_init=0.0001 |  |
|  | 'alpha': np.logspace(-4, 4, 10) | L2 penalty (regularization term) |

Table S3. ChatGPT response using GPT-4 backend to identify causal directions of the correlations.

| Variable 1 | Variable 2 | Response |
| --- | --- | --- |
| Age | DM duration | To analyze which cause-and-effect relationship is more plausible between changing age and changing the duration of diabetes mellitus (dm_duration), let's consider what each variable represents and the nature of causation. 1. **Understanding Age and dm_duration**: - **Age**: This is the participant-reported age at the time of the survey. Age is a straightforward variable that increases as time progresses. - **dm_duration**: This measures how long a participant has had diabetes mellitus, based on medical records from the past three months. This duration increases as time since diagnosis progresses. 2. **Analyzing Causal Direction**: - **Option A: Changing Age Causes a Change in dm_duration**: - This scenario implies that as a person gets older, the duration since they were diagnosed with diabetes mellitus (if they have it) also increases. This is a logical outcome because the start point of measuring dm_duration is the time of diabetes diagnosis, which remains fixed, and as time moves on (age increases), the duration naturally increases. - **Option B: Changing dm_duration Causes a Change in Age**: - This scenario would imply that increasing the duration of having diabetes would somehow influence how old a person is. This is illogical because the flow of time (and thus aging) cannot be influenced by the duration of a medical condition. 3. **Logical Assessment**: - Given that age progresses naturally over time and that the measurement of dm_duration starts from a fixed point (diagnosis), it is clear that age influences dm_duration simply through the passage of time. The reverse—dm_duration influencing age—is not feasible as the progression of age is a continuous, inevitable process that cannot be altered by changes in health conditions. Based on this reasoning, the cause-and-effect relationship that is more likely is: **A**. Changing age causes a change in dm_duration. Final answer: **A** |
| Age | Marital status | To determine which cause-and-effect relationship is more likely between the two provided options (A and B), we need to consider how each variable could influence the other based on their definitions and general understanding: ### Step-by-step Analysis: 1. **Understanding the Variables**: - **Age**: This is a measure of how many years a person has lived, which naturally increases as time passes. - **Marital Status**: This is a classification of a person's relationship status, categorized into unmarried, married, widowed, or divorced. 2. **Direction of Causality**: - **Option A: Changing Age Causes a Change in Marital Status**: - As people age, they typically undergo various life stages and experiences. Younger individuals may start unmarried, older individuals might more commonly be married, and even older age groups could increasingly be widowed or divorced. - This suggests that as individuals age, it's natural for their marital status to change in response to life events that are often age-related (e.g., deciding to marry, experiencing the death of a spouse, or divorcing). - **Option B: Changing Marital Status Causes a Change in Age**: - Marital status is a state that can change multiple times throughout a person's life, but these changes are independent of the person's biological age. For instance, getting married or divorced does not alter a person's age. - Age is a chronological measurement and cannot be influenced by marital status. No matter the change in marital status, the biological age will continue to increase uniformly. 3. **Logical Deduction**: - It is clear that age is a sequential and continuous increase that everyone experiences, and it cannot be influenced by marital status or almost any other external social factor. - Conversely, changes in life stages, which are often age-related, can influence changes in marital status. For instance, it's common to marry as one grows older from adolescence into early adulthood, or to become widowed as one ages into the later stages of life. ### Conclusion: Given the logical flow and understanding of how these variables interact based on typical life progressions, it is more plausible that age influences marital status rather than the reverse. Final answer: **A**. |
| Sex | Cigarrette smoking | To determine which cause-and-effect relationship is more likely between changing sex and changing cigarette smoking habits, let's consider each scenario step-by-step based on definitions and general understanding: 1. **Understanding of Biological Sex**: - Biological sex (female or male) is typically a genetically determined characteristic established at birth based on physiological attributes such as chromosomes, hormone levels, and reproductive organs. - While individuals may undergo sex change procedures (gender reassignment surgery and hormone treatments) to transition to the gender they identify with, this is a complex process that involves medical, psychological, and social changes, and is related to gender identity rather than the binary biological categorization of sex. 2. **Understanding of Cigarette Smoking**: - Cigarette smoking is a behavior that involves the inhalation of smoke from burning tobacco encased in cigarettes. - Smoking status can change over time based on personal habits, health considerations, social influences, and access to resources. 3. **Analyzing Cause-and-Effect**: - **Scenario A: Changing Sex Causes a Change in Cigarette Smoking**: - Changes in sex, as defined by biological attributes, are relatively fixed and not subject to frequent change. However, the transition processes involved in changing gender identity could influence lifestyle choices, including smoking, due to stress, social dynamics, or personal health decisions. This implies a potential, albeit indirect, impact on smoking behavior due to changes in gender-related life circumstances. - **Scenario B: Changing Cigarette Smoking Causes a Change in Sex**: - The act of smoking or quitting smoking is unlikely to have any direct physiological impact on one's biological sex. Changes in smoking habits can affect health but do not alter genetic or primary biological characteristics that define sex. Given these analyses, it's evident that the causal connection in Scenario A, while indirect and more related to gender identity than biological sex, is more plausible than Scenario B, where the behavior of smoking has no biological mechanism to alter sex. **Final Answer:** A |
| Comorbidity | Cigarrette smoking | To determine which cause-and-effect relationship is more likely between changing comorbidity status and changing cigarette smoking behavior, we can use the following reasoning: ### Step-by-Step Analysis 1. **Definition Understanding**: - **Comorbidity**: This refers to the presence of one or more additional medical conditions co-occurring with a primary condition. This is determined by reviewing medical records. - **Cig_smoking**: This is based on self-reported data on whether an individual smokes cigarettes at the time of the survey. 2. **Direction of Causality**: - **A. Comorbidity → Cig_smoking**: This suggests that changes in the health status (i.e., gaining or losing a comorbid condition) could influence an individual's smoking habits. For example, a diagnosis might lead to lifestyle changes due to health concerns or recommendations from a health professional. - **B. Cig_smoking → Comorbidity**: This implies that smoking could lead to changes in health status, potentially causing or exacerbating comorbid conditions. Smoking is well-documented to increase the risk of various diseases, including respiratory and cardiovascular conditions. 3. **Evaluating Likelihood of Each Scenario**: - **Scenario A**: - While it's plausible that a change in health status could influence an individual to start or stop smoking (e.g., a new health diagnosis prompting cessation), the direct impact of comorbidity on smoking behavior is less straightforward without considering psychological, social, and behavioral factors. - **Scenario B**: - There is extensive evidence supporting the fact that smoking can cause or worsen many health conditions, thereby increasing the likelihood or severity of comorbidities. This pathway is well established in medical literature. 4. **Conclusion**: - Based on the directness and strength of evidence, the relationship where smoking affects health (and thus potentially increases comorbidities) is more likely and straightforward compared to the scenario where comorbidities would lead someone to change their smoking habits. The causal path from smoking to increased health risks is well documented and biologically plausible. ### Final Answer <B> |
| Comorbidity | HbA1c | To determine which cause-and-effect relationship is more likely between changing comorbidity and changing HbA1c levels, we need to consider the definitions and understand the nature of each variable: 1. **Comorbidity**: This refers to the presence of one or more additional medical conditions co-occurring with a primary condition. Comorbidity in this context is a categorical variable (yes/no) based on medical records from the past three months. 2. **HbA1c (Glycated Hemoglobin)**: This is a measure of the average blood glucose concentration over the past three months. It reflects the long-term management of blood glucose levels and is a continuous variable expressed in percentage. ### Causal Reasoning Consideration: - **Direction from HbA1c to Comorbidity (Option B)**: - **Biological Plausibility**: Long-term elevated blood glucose levels (high HbA1c) can lead to a range of complications and comorbid conditions like cardiovascular diseases, neuropathy, retinopathy, and kidney disease. Thus, there is a biologically plausible pathway where poor blood glucose control could lead to an increase in comorbid conditions. - **Temporal Precedence**: HbA1c reflects blood glucose control over approximately three months and can precede the onset or worsening of comorbid conditions that are influenced by glucose levels. - **Direction from Comorbidity to HbA1c (Option A)**: - **Biological Plausibility**: While comorbid conditions can affect overall health status, the direct influence of a comorbid condition on HbA1c levels is less clear unless the specific comorbidities directly relate to glucose metabolism (e.g., pancreatic diseases). However, in a general context where any comorbidity (not specifically related to glucose metabolism) could affect HbA1c, the causal pathway is less direct. - **Temporal Precedence**: Comorbid conditions might influence overall health management, including lifestyle and treatment adherence, which can indirectly influence HbA1c levels. However, this influence is generally less direct compared to the impact of HbA1c on developing new comorbid conditions. ### Conclusion: Considering the biological plausibility and the temporal precedence, it is more biologically plausible and logically consistent that poor control of blood glucose levels (as indicated by HbA1c) would lead to an increase in the likelihood or severity of comorbid conditions. Thus, the causal relationship from HbA1c to comorbidity seems more likely due to its direct impact on health outcomes related to chronic high blood sugar. **Final Answer: B** |
| Comorbidity | Fatigue | To approach the question of which cause-and-effect relationship is more likely, we'll analyze the definitions and the logical implications of each scenario. **Definitions and Implications:** 1. **Comorbidity**: This is defined based on medical records over the past three months, indicating a presence (yes) or absence (no) of additional diseases or conditions alongside a primary condition. 2. **Fatigue**: This is based on participant-reported feelings at the time of the survey, indicating whether they feel fatigued (yes) or not (no). **Cause-and-Effect Analysis:** - **Scenario A: Changing Comorbidity Causes a Change in Fatigue** - **Logical Path**: The presence of comorbidities (additional health issues) could logically lead to changes in an individual's overall health status, including increased feelings of fatigue. Comorbidities often complicate health management and could directly impact energy levels and recovery processes, leading to fatigue. - **Medical Perspective**: Many health conditions that might be considered comorbidities, such as diabetes, heart disease, or chronic pain, are known to influence fatigue levels due to their systemic effects on the body. - **Scenario B: Changing Fatigue Causes a Change in Comorbidity** - **Logical Path**: While fatigue can be a symptom of many medical conditions, it is less likely to be a causative factor for the presence of comorbidities. Fatigue itself is a symptom rather than a disease or condition, and while it can exacerbate existing conditions, it does not typically cause new comorbid conditions to arise. - **Medical Perspective**: Fatigue could influence the perception of health or exacerbate symptoms, but it does not create comorbid conditions; rather, it might lead to investigations that could uncover pre-existing comorbidities. **Conclusion:** Considering the implications of each scenario, it is more plausible that the presence of comorbidities would influence the level of fatigue rather than fatigue causing comorbidities. Comorbid conditions often have significant impacts on overall health, which can manifest as increased fatigue due to the body's heightened strain from managing multiple health issues. Therefore, the more likely cause-and-effect relationship is: <**A**> |
| OHA + insulin | Cigarrette smoking | To determine which cause-and-effect relationship is more likely between changing dm_treatment and changing cig_smoking, we'll analyze the plausibility and directionality of the potential effects between these variables. ### Step 1: Understanding the Variables 1. **dm_treatment**: This involves the type of treatment a patient receives for diabetes mellitus, including oral hypoglycemic agents or a combination with insulin. This is influenced by the patient’s health status and medical needs regarding their diabetes control. 2. **cig_smoking**: This refers to whether an individual currently smokes cigarettes or not. This is a lifestyle choice and could have various health implications. ### Step 2: Analyzing the Direction of Influence #### A. Changing dm_treatment causes a change in cig_smoking - **Plausibility**: This implies that modifying the treatment for diabetes (like prescribing a different medication) would directly influence a patient's smoking habits. - **Analysis**: While changes in medication can impact a patient's overall health and may indirectly affect lifestyle choices due to changes in health status or doctor's advice, it's less likely that such changes would directly cause a patient to start or stop smoking. Medical treatment primarily targets the disease condition, not lifestyle habits directly, unless specifically designed as part of a broader intervention program that includes lifestyle coaching. #### B. Changing cig_smoking causes a change in dm_treatment - **Plausibility**: If a patient changes their smoking habits, this could affect their overall health, particularly in the context of diabetes. Smoking has been shown to exacerbate many health conditions, including cardiovascular diseases and complications in diabetes management. - **Analysis**: If a patient stops smoking, their overall health might improve, potentially leading to changes in how their diabetes is managed, such as adjusting dosages or types of medication due to improved health metrics or reduced risk factors. Conversely, if a patient starts or increases smoking, their diabetes may become harder to manage, necessitating a more aggressive treatment approach. ### Step 3: Conclusion Given these considerations, it seems more plausible and direct that changes in smoking habits could influence the management and treatment plan of diabetes rather than changes in diabetes treatment directly influencing smoking habits. The lifestyle choice of smoking directly impacts health and can influence the severity or management of chronic conditions like diabetes. Therefore, the more likely cause-and-effect relationship is: <B>B. changing cig_smoking causes a change in dm_treatment</B>. |
| HbA1c | Fatigue | To address the question regarding the cause-and-effect relationship between changing HbA1c levels and fatigue, we need to consider the biological and physiological mechanisms that might support each direction of causality. ### Step 1: Understanding HbA1c HbA1c, or glycated hemoglobin, is a measure of the average blood glucose concentration over the past three months. It is commonly used to diagnose and monitor diabetes. High levels of HbA1c indicate poorer blood sugar control and can reflect an increased risk of diabetes-related complications. ### Step 2: Understanding Fatigue Fatigue in this context is subjectively reported by participants and can be influenced by various factors including physical health, mental health, sleep quality, and metabolic states. ### Step 3: Analyzing Causality #### A. Changing HbA1c Causes a Change in Fatigue - **Biological Plausibility**: Poor blood sugar control as indicated by elevated HbA1c can lead to metabolic imbalances that might result in fatigue. High blood glucose levels can cause dehydration (due to frequent urination) and metabolic disturbances, which can lead to fatigue. - **Mechanism**: The metabolic stress from fluctuating or consistently high blood sugar could directly affect cellular energy production, leading to feelings of tiredness or exhaustion. #### B. Changing Fatigue Causes a Change in HbA1c - **Biological Plausibility**: While fatigue can affect a person’s lifestyle choices, such as physical activity and diet, which in turn can influence blood glucose levels and potentially HbA1c, the direct impact of fatigue on glycated hemoglobin levels is less straightforward. - **Mechanism**: If fatigue leads to reduced physical activity, there might be an indirect effect on blood glucose control. However, the direct biological pathway through which fatigue itself would alter glycated hemoglobin levels is less clear. Fatigue does not chemically or physiologically alter HbA1c but may influence behaviors that could indirectly affect HbA1c levels over a longer period. ### Conclusion Based on the physiological links and the directness of the impact, it is more likely that changes in HbA1c levels cause changes in fatigue rather than the reverse. The pathway from poor blood sugar control to fatigue is direct and well-supported by metabolic biology, whereas the pathway from fatigue to changes in HbA1c is more indirect and depends on behavioral changes rather than physiological changes in blood chemistry. **Final Answer: <A>** |
| Sex | OHA + insulin | To determine which cause-and-effect relationship is more plausible between changing sex and changing diabetes mellitus treatment (dm_treatment), let’s consider what each variable represents and how changes in these variables can reasonably be expected to occur based on their definitions. ### Step 1: Understanding the Variables 1. **Sex**: This variable is defined as the participant-reported biological sex (female or male). Generally, biological sex is considered a stable characteristic that does not change frequently. Changes in biological sex as recorded in medical or other formal contexts could involve legal or medical alterations such as those observed in transgender individuals undergoing sex reassignment. 2. **Dm_treatment**: This represents the type of treatment an individual receives for diabetes mellitus, categorized as oral hypoglycemic agents or a combination of oral hypoglycemic agents and insulin. This treatment is typically selected based on clinical factors including the severity of diabetes, effectiveness of current treatment, and patient health status. ### Step 2: Evaluating Causality Directions **A. Changing sex causes a change in dm_treatment**: - The direction implied here is that a change in the participant’s biological sex could lead to a change in how diabetes is treated. While there are scenarios where hormonal therapy and other treatments associated with sex reassignment could necessitate adjustments in various medical treatments (including potentially diabetes treatment due to changes in hormone levels and body composition), such cases are specific and less common. **B. Changing dm_treatment causes a change in sex**: - This suggests that modifying the type of diabetes treatment would cause a change in the biological sex of the individual. Medically and biologically, this is not feasible. Treatments for diabetes, including oral medications and insulin, do not influence or change an individual's biological sex. ### Step 3: Drawing Conclusions Given the definitions and the typical medical understanding of these variables: - **Option A** might be plausible in rare and specific medical contexts where sex reassignment and related treatments could impact the broader management of chronic conditions like diabetes. - **Option B** is implausible as dm_treatment does not and cannot cause changes in a person's biological sex. ### Final Decision Considering the explanations above, the more likely and medically reasonable cause-and-effect relationship is: <**A**> |
| OHA + insulin | Fatigue | To determine which cause-and-effect relationship is more likely, we can analyze the definitions and the logic behind how one variable could influence the other. Let’s break down each relationship step by step: ### A. Changing dm_treatment causes a change in fatigue 1. **Understanding dm_treatment**: This variable represents the type of diabetes mellitus treatment the patient is undergoing. This could include an oral hypoglycemic agent, a combination of oral hypoglycemic agents, or a combination of oral hypoglycemic agents and insulin. Changes in treatment can occur due to a variety of reasons such as changes in disease status, side effects, or doctor’s advice. 2. **Potential Impact on Fatigue**: Different diabetes treatments might have varying side effects and impacts on a patient's metabolic state. For example, some treatments might affect blood sugar levels more dramatically than others. Since fatigue in diabetes patients can be linked to blood sugar fluctuations, changing the treatment might lead to a change in fatigue levels, either increasing or decreasing them. ### B. Changing fatigue causes a change in dm_treatment 1. **Understanding Fatigue**: Fatigue here is measured based on patient reports at the time of the survey. It indicates a subjective feeling of tiredness, which can be influenced by various factors, including diabetes management, other health conditions, sleep quality, or physical activity. 2. **Potential Impact on dm_treatment**: If a patient reports increased fatigue, this might prompt a healthcare provider to review their current diabetes management strategy, including treatment. The idea here is that if a patient feels more fatigued, it might be a sign that the current treatment is not optimal or needs adjustment. However, it's less direct than the effect of treatment on fatigue because fatigue can be caused by many factors not necessarily related to diabetes treatment directly. ### Logical Causality and Medical Understanding - **Direct Influence**: Changing dm_treatment has a more direct influence on fatigue because medical treatments are designed to alter physiological states directly, and these changes can lead to side effects like fatigue. - **Indirect Influence**: Changes in fatigue influencing dm_treatment is more of an indirect pathway, where the change in symptoms might prompt a reconsideration of treatment, but it’s not a guarantee that treatment will change. Moreover, fatigue could prompt a review of many aspects of health, not just dm_treatment. Given these analyses, the more likely cause-and-effect relationship based on direct influence and specific targeting in treatment is: **A. Changing dm_treatment causes a change in fatigue.** So, the final answer within the tags is: **A** |

DM, diabetes mellitus; OHA, oral hypoglycemic agent.

Table S4. Regression analysis for SCM.

| Variable |  | Covariates | | OR (95% CI; p-value) |
| --- | --- | --- | --- | --- |
|  |  | Confounder | Mediator |  |
| HbA1c | 6.5+ vs. 5.7 to <6.5 | - | - | 5.7 (2.7, 12.2; <0.001) |
|  |  | - | Comorbidity | 6.1 (2.5, 14.4; <0.001)* |
| Comorbidity | yes vs. no | - | - | 24.6 (12.1, 50.0; <0.001) |
|  |  | HbA1c | - | 25.2 (12.1, 52.4; <0.001)* |
| OHA + insulin | OHA + insulin vs. OHA | - | - | 3.6 (2.0, 6.4; <0.001) |

*, adjusted OR; OHA, oral hypoglycemic agent; OR, odds ratio; SCM, structural causal modeling.
